# Hospital-treated infectious diseases and the risk of dementia: multicohort study with replication in the UK Biobank

**DOI:** 10.1101/2020.04.20.20072355

**Authors:** Pyry N Sipilä, Nelli Heikkilä, Joni V Lindbohm, Christian Hakulinen, Jussi Vahtera, Marko Elovainio, Sakari Suominen, Ari Väänänen, Aki Koskinen, Solja T Nyberg, Jaana Pentti, Timo E. Strandberg, Mika Kivimäki

## Abstract

**Background:** Infectious diseases have been hypothesised to increase the risk of dementia. However, the evidence is sparse, captures only a limited range of infectious diseases, and relies on short follow-up periods. We assessed a wide range of severe (hospital-treated) bacterial and viral infections and their subtypes as risk factors for dementia in three large cohorts followed up for almost two decades and replicated the main findings in the UK Biobank.

**Methods:** For primary analysis, we pooled individual-level data from three prospective cohort studies with a median follow up of 19 years (from 1986-2005 to 2012-2016) and a total of 273 125 dementia-free community-dwelling participants. The replication analysis with the UK Biobank was based on 492 146 individuals (median follow-up 9·0 years from 2006-2010 to 2018). We ascertained exposure to infectious diseases and their subtypes before dementia onset using linked records from national hospital inpatient registers. Incident dementia was identified from linked hospital inpatient and outpatient records, medication reimbursement entitlements, and death certificates.

**Findings:** In the primary analysis based on 5·3 million person-years at risk, 88 099 participants had a hospital-treated infection before dementia onset and 3064 developed dementia. Gram-negative bacterial infections (hazard ratio [HR] 1·64; 95% confidence interval [CI] 1·25–2·14) and herpesvirus infections (HR 1·96; 95% CI 1·31–2·93) were robustly associated with an increased risk of dementia. For these infections, the relative risk of dementia remained similar when reverse causation and ascertainment biases were minimised by assessing only new dementia cases that occurred more than 10 years after the infection, and when comorbidities and potential confounders were considered. The associations were replicated in the UK Biobank with stronger relations observed for vascular dementia than Alzheimer’s disease. In contrast to gram-negative bacterial and herpesvirus infections, the hazard ratio for all bacterial infections combined attenuated from 1·60 (95% CI 1·48–1·72) to 1·27 (95% CI 1·14–1·41) when bias was minimised. For all viral infections, the corresponding attenuation was from HR 1·63 (95% CI 1·32–2·00) to 1·19 (95% CI 0·87–1·62).

**Interpretation:** Gram-negative bacterial infections and herpesvirus infections were associated with a moderately increased risk of dementia both in the short- and long-term. Hospital-treated bacterial and viral infections in general had only modest long-term associations with incident dementia.

**Funding:** UK Medical Research Council, US National Institute on Aging, NordForsk, Academy of Finland, Helsinki Institute of Life Science.

**Research in context:** *Evidence before this study:* Infectious diseases have been hypothesised in the aetiology of dementia. We searched PubMed on 2 April 2020 for observational studies and systematic reviews using the search terms ((Alzheimer* OR dementia) AND infectio* AND ((systematic[sb]) OR (Observational Study[ptyp])) without restrictions on language or publication date. We also searched the reference lists and citations of relevant articles. In observational studies, infectious diseases in general and specific bacterial (sepsis, septicaemia, bacteraemia, pneumonia, osteomyelitis, urinary tract infection, cellulitis, syphilis, and *Borrelia burgdorferi* and *Chlamydia pneumoniae* infections) and viral infections (hepatitis C infection, HIV) are linked to an increased risk of dementia. Additionally, there was suggestive evidence for associations of herpesvirus infections, *Toxoplasma gondii* parasite infection, and poor oral health with dementia. However, no large-scale studies assessed a wide range of different types of infectious diseases systematically in a single analytical setting and with adequate control for potential sources of bias, such as reverse causation resulting from the systemic changes related to the preclinical stages of dementia that increase susceptibility to infectious diseases.

*Added value of this study:* In this multicohort study, the primary analysis was based on individual-level data from 273 125 dementia-free community-dwelling participants with a median follow-up of 19 years. We ascertained 528 different infectious diseases before dementia onset using national hospital inpatient records and categorised them by causative microorganism (bacteria, viruses), disease invasiveness (invasive vs localised bacterial infections), and subtype (extracellular vs intracellular lifestyle and cell wall structure of bacteria, and type of virus). When reverse causation was minimised by analysing only infections that occurred more than 10 years before dementia onset, the relative risk of dementia was modest for all bacterial infections combined (hazard ratio [HR] 1·27; 95% confidence interval [CI] 1·14–1·41) and for all viral infections combined (HR 1·19; 95% CI 0·87–1·62). The associations of bacterial infections with dementia did not differ by the invasiveness of the infection (invasive vs localised infections) or extracellular vs intracellular lifestyle of the causative microorganism. In analysis of specific diagnostic groups, robust and stronger long-term associations with dementia were observed for Gram-negative bacterial infections (HR 1·64, 95% CI 1·25–2·14 for full follow-up; HR 1·67, 95% CI 1·13–2·48 from year 10 onwards) and herpesvirus infections (HR 1·96, 95% CI 1.31–2.93 full follow-up; HR 1·92, 95% CI 1·11–3·32 from year 10 onwards). These associations remained after controlling for comorbidities and potential confounders and were replicated in an independent cohort of 492 146 individuals from the UK Biobank (median follow-up 9·0 years) with stronger relations observed for vascular dementia than Alzheimer’s disease.

*Implications of all the available evidence:* Gram-negative bacterial infections and herpesvirus infections seem to be robust long-term risk factors for dementia. Further mechanistic and molecular research is warranted to clarify the potential role of Gram-negative bacterial infections and herpesvirus in neurodegeneration and the aetiology of dementia.

## Introduction

Several lines of research suggest a role for inflammation and infectious diseases in the aetiology of dementia.^1–7^ Many genes that predispose to dementia are involved in inflammatory pathways.^1^ Systemic inflammation is also associated with accelerated cognitive decline and dementia in prospective studies.^8–10^ In addition, associations between infectious diseases and dementia have been found in several independent cohorts.^11–13^

There are plausible mechanisms linking systemic inflammation and infectious diseases to the development of dementia. Microglia, the resident macrophages of the central nervous system, play an important role in neuroinflammation and neurodegeneration.^1^ A recent study suggests that peripheral inflammatory stimuli, such as lipopolysaccharides from Gram-negative bacteria, can prime microglia to a proinflammatory state that may persist long after the initial insult and potentially contribute to the development of Alzheimer’s pathology.^2^ Microbes may also trigger autoimmunity in the central nervous system through molecular mimicry^14^ and some intracellular bacteria may directly infect the central nervous system.^5^ Moreover, β-amyloid itself may be an antimicrobial peptide such that infectious inflammatory stimuli partly drive amyloidosis in the brain.^15^

However, there are important gaps in the present knowledge. Observational evidence is scarce, relies on short follow-up periods, and captures only a limited range of infectious diseases.^11–13,16^ Given the long preclinical phase of dementia that lasts years or decades, the associations between infectious diseases and dementia in these studies may be inflated by reverse causation (i.e., by increased susceptibility to infections due to systemic changes related to preclinical dementia) or ascertainment bias (a diagnosis of infection may increase the likelihood of diagnosing other conditions of the patient, including dementia).^6,11^ The mechanistic studies on infection, neuroinflammation, and neuropathology, in turn, lack external validity as they are mostly from in vitro or animal models; the few studies in humans cover a limited range of infections (sepsis, pneumonia, osteomyelitis, cellulitis, urinary tract, and herpesvirus infections).^3,4,11–13,16^ Due to these limitations it is difficult to determine whether the observed associations with dementia are attributable to infections in general or only to certain types of infections, or represent artefacts.

We performed a large-scale study to systematically assess the long-term dementia risk associated with a wide range of different types of infectious diseases. To capture the typical nature of the inflammatory response associated with each disease,^17^ we classified infectious diseases by causative microorganism (bacteria, viruses), disease invasiveness (invasive vs localised bacterial infections), and subtype (extracellular vs intracellular lifestyle and cell wall structure of bacteria, and type of virus). To obtain reproducible findings, we repeated the main analyses in a large independent study population.

## Methods

### Study population

The primary analysis of this study was based on pooled individual-level data from three harmonised prospective cohort studies linked to national health registries in Finland: the Finnish Public Sector study (FPS), the Health and Social Support study (HeSSup), and the Still Working study (STW). Of the 337 120 eligible participants, we included 273 125 individuals aged ≥18 years and free of known dementia when they entered the study (baseline 1986-2005). Data collection and analysis were approved by the ethics committee of the Helsinki and Uusimaa Hospital District (FPS), the Turku University Central Hospital Ethics Committee (HeSSup), and the ethics committee of the Finnish Institute of Occupational Health (STW). References, details, and a flowchart describing the selection of participants are provided in appendix (pp. 2-6).

We used the UK Biobank as an independent replication cohort (https://www.ukbiobank.ac.uk/). Using the same criteria as in the primary analysis, a total of 492 146 dementia-free individuals were included in the analysis (baseline 2006-2010). This study was performed under a generic approval from the National Health Service National Research Ethics Service (June 17, 2011; Ref 11/NW/0382).

### Exposure to hospital-treated infectious diseases

We defined and classified hospital-treated infectious diseases according to the International Classification of Diseases, 10th Revision (ICD-10). The diseases were classified hierarchically to reflect their likely severity and the type of immune response associated with different pathogens (**Figure 1**).^17^

**Figure 1.**
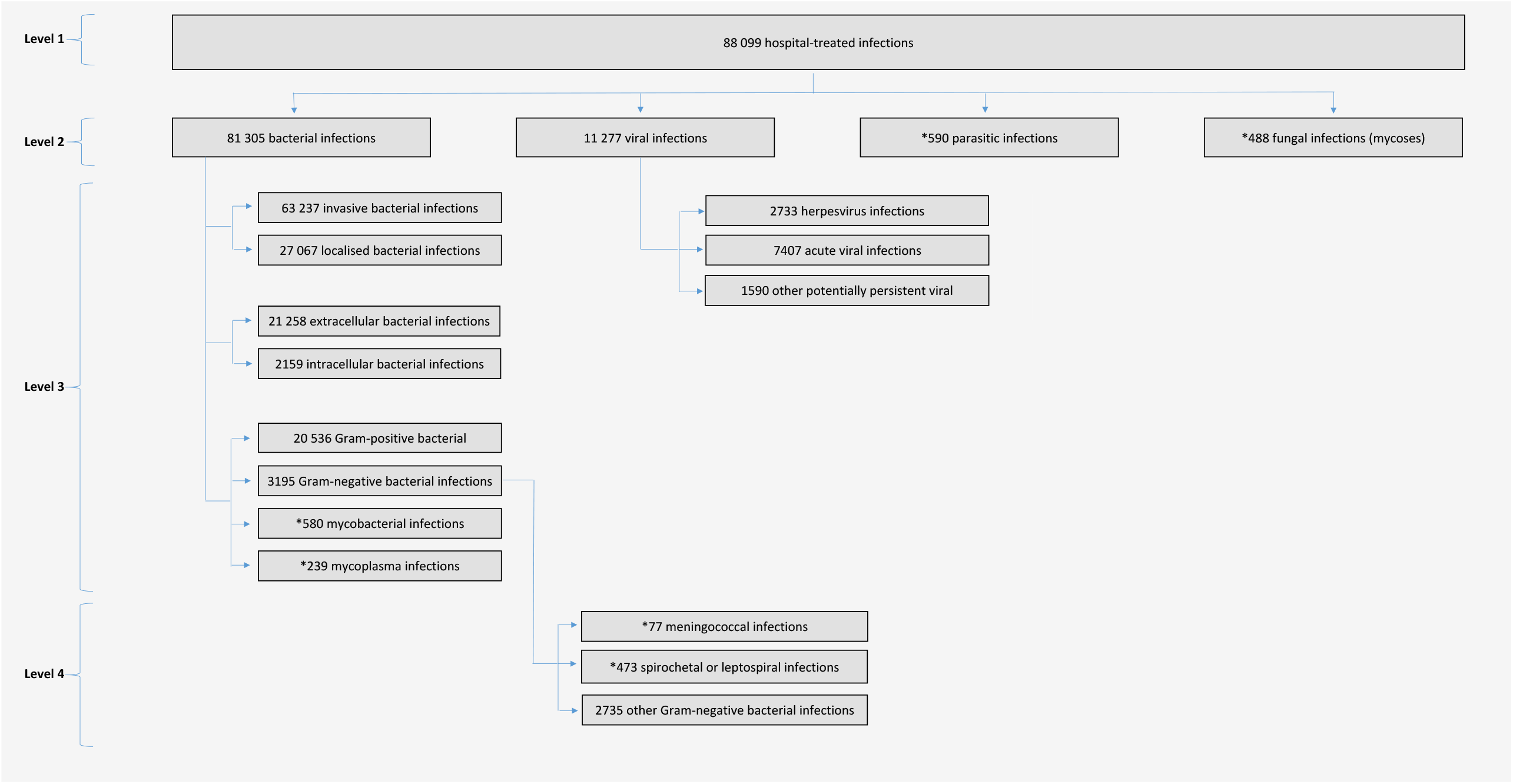
Classification of hospital-treated infectious diseases in the study and number of cases in the primary analysis. Note: The numbers of different infectious diseases add up above the total number of infections, because some participants have been hospitalised for more than one infectious disease. *Too rare to be analysed separately.

Level 1 consisted of all infectious diseases. On level 2, we divided infectious diseases into bacterial, viral, parasitic, or fungal. On level 3, we classified bacterial infections further according to disease invasiveness ([potentially] invasive vs [mostly] localised bacterial infections), bacterial lifestyle (extracellular vs obligate or facultative intracellular bacteria), and cell wall structure (Gram-positive bacteria vs Gram-negative bacteria vs mycobacteria vs mycoplasma). All classifications of bacterial lifestyle and cell wall structure were based only on those ICD-10 codes that defined the causative microorganism unambiguously (e.g., shigellosis, legionnaires disease, pneumonia due to *Haemophilus influenzae*). Invasive bacterial infections included, for example, typhoid fever, other salmonella infections, tuberculosis, sepsis, erysipelas, bacterial meningitis, pneumonia, osteomyelitis, and pyelonephritis. Localised bacterial infections included, for example, gastroenteritis, otitis externa and media, acute sinusitis, pharyngitis, tonsillitis, laryngitis and bronchitis, dental caries, periodontitis, mild skin infections, acute cystitis, and localised puerperal infections. A full list of invasive and localised bacterial infections is provided in appendix (pp. 31-37).

On level 3, we also classified viral infections to acute infections that are typically eradicated by the immune system; herpesvirus infections that persist in the body after primary infection; and other potentially persistent viral infections (such as hepatitis B and C and HIV). On level 4, we further divided Gram-negative bacterial infections to those with known neurological sequelae (meningococcal infections, spirochaetal and leptospiral infections)^18,19^ and other Gram-negative infections. The appendix (pp. 14–51) describes the definitions and ICD-10 codes of all the disease categories.

We linked participants to the records of national health registers and retrieved both primary and secondary diagnoses of the infectious diseases from inpatient hospital discharge information using ICD-10 codes. Diagnosis codes from the 8th and 9th revisions (ICD-8 and ICD-9) were converted into the corresponding ICD-10 codes. The appendix (pp. 52-75) describes the ICD-10 codes in the study with their corresponding ICD-8 and ICD-9 codes.

### Ascertainment of incident dementia after exposure to infection

We retrieved diagnoses of incident dementia from four sources, including hospital inpatient records; medication reimbursement entitlements for the treatment of dementia (these require a diagnosis of Alzheimer’s disease or Parkinson’s disease dementia made in a neurology or geriatrics unit in secondary or tertiary health care or by a qualified neurologist or geriatrician); causes of death; and, for FPS and SWT, hospital outpatient records (appendix pp. 3-6). The first occurrence of dementia diagnosis, whether primary or secondary, in any of these information sources defined the date of incident dementia.

A diagnosis of all-cause dementia consisted of ICD-10 codes F00-F03, F05·1, G30, G31·0, G31·1, G31·8, and the corresponding ICD-8 (29000-29019, 34791, 34792) and ICD-9 (290, 2900A, 2941A, 3310A, 3311A, 3312X, 4378A) codes in the Finnish national editions of the International Classification of Diseases. The subtypes of dementia were defined as follows: Alzheimer’s disease (F00, G30, 29010, 3310A), frontotemporal dementia (G31·0, F02·0, 29011, 34791, 3311A), Parkinson’s disease dementia (F02.3), vascular dementia (F01, 4378A), other defined dementia (G31·8, F02·1, F02·2, F02·4, F02·8), or unspecified dementia (F03, G31·1, F05·1, F02·39, 29000, 29019, 290, 2900A, 2941A, 34792, 3312X). In the UK Biobank, we defined all-cause dementia and its subtypes using the validated cohort algorithms. For dementia subtypes with no algorithm available, we used diagnoses from hospital admission records (appendix pp. 5-6).

### Assessment of covariates and comorbidities

The covariates at study entry were chosen to reflect the following potential common risk factors for infections and dementia:^20–22^ sex, socioeconomic status (low, intermediate, high), smoking (never smokers, ex-smokers, current smokers), and alcohol drinking (non-drinkers, moderate drinkers, intermediate drinkers, heavy drinkers). In the UK Biobank, we additionally included body mass index measured at baseline. Socioeconomic status was measured by education (FPS, HeSSup, UK Biobank) or occupational grade (STW).

Hypertension, diabetes mellitus, ischaemic heart disease, cerebrovascular disease, and Parkinson’s disease are considered as comorbidities that are potential common risk factors for infections and dementia. These comorbidities were measured using primary and secondary diagnoses from hospital inpatient discharge information and, as available, prescription medication reimbursement entitlements at baseline and during follow up. In the UK Biobank, we additionally used information from measurements and self-reports.

In appendix (pp. 3-6 and 76-77), the exact definitions and distributions of the covariates and comorbidities in each cohort are provided.

### Statistical analysis

Exposure to infections was modelled using time-dependent analysis.^23^ Participants with infection at baseline were considered exposed. Those with infection during follow up were considered non-exposed until the onset of infection (date of hospitalisation) and exposed thereafter. Participants who did not have infection at baseline or follow up were considered unexposed across the entire follow up. Follow up for incident dementia started from study entry (that is, the date of return of baseline questionnaire, or the year of identification from the employer’s register when no questionnaire data were used) and spanned until first dementia diagnosis, death, or end of follow up (2012-2016 in primary cohorts, 2018 in replication cohort), whichever came first.

We used Cox proportional hazards models to compute hazard ratios (HR) for the associations of each infectious disease or disease group (index infection) with incident dementia. In the primary analysis, we pooled individual-level data from three cohort studies and took the within-study clustering of participants into account using cohort-specific baseline hazards and cohort-specific adjustment terms for covariates.^24^ We adjusted the analyses for sex and socioeconomic status and used age as the time scale. The proportional hazards assumption was examined using scaled Schoenfeld residuals (appendix pp. 7-10).

To reduce the risk of reverse causation and ascertainment bias, we repeated the analysis after excluding incident dementia cases that occurred during the first 10 years after initial hospitalisation due to the index infection. For those not exposed to the index infection by study entry, we also excluded dementia cases occurring within the first 10 years of follow up (appendix pp. 11–12). In other sensitivity analyses, we repeated the main analyses after exclusion of participants with diabetes, cardio- or cerebrovascular disease, or Parkinson’s disease and adjusted analyses for heavy drinking, smoking, and body mass index at study entry. We repeated analyses of the main findings in the UK Biobank.

Data analysis was performed using Stata MP 15 and 16. The syntax for the analyses is available in the appendix (pp. 79-112). All confidence intervals (CI) are reported on a 95% confidence level.

## Results

Table 1 describes the participants’ demographic characteristics and the appendix (pp. 13) shows the age distribution at incident dementia. In the primary analysis, 88 099 participants were hospitalised for an infection. During 5 306 852 person-years at risk, we identified 3064 incident cases of dementia. Of these, 1934 were diagnosed as Alzheimer’s disease, 219 as vascular dementia, 5 as comorbid Alzheimer’s disease and vascular dementia, and 906 as other or unspecified dementias. By source of identification, we identified 1149 incident dementia cases from inpatient hospital discharge records, 814 from other hospital records, 1076 from anti-dementia medication reimbursement entitlements, and 25 from death certificates.

**Table 1.** Demographic characteristics of participants at study entry in primary and replication cohorts.

|  | **Pooled data from 3 cohorts**<br>**(N = 273 125)** | **UK Biobank**<br>**(N = 492 146)** |
| --- | --- | --- |
| Age at entry, years |  |  |
| 18-39 | 181 773 (66.6%) | 4 (0.0%) |
| 40-49 | 57 850 (21.2%) | 114 969 (23.4%) |
| 50-59 | 31 558 (11.6%) | 163 758 (33.3%) |
| 60-87 | 1944 (0.7%) | 213 415 (43.4%) |
| Sex |  |  |
| Men | 80 765 (29.6%) | 267 940 (54.4%) |
| Women | 192 360 (70.4%) | 224 206 (45.6%) |
| Socioeconomic status |  |  |
| Low | 41 803 (15.3%) | 85 203 (17.3%) |
| Intermediate | 93 191 (34.1%) | 245 836 (50.0%) |
| High | 138 131 (50.5%) | 161 107 (32.7%) |
| Follow-up, years |  |  |
| Median (range) | 19.0 (0.0-30.8) | 9.0 (0.0-11.1) |
| Dementia by the end of follow-up |  |  |
| No | 270 061 (98.9%) | 489 771 (99.5%) |
| Yes | 3064 (1.1%) | 2375 (0.5%) |
| Age at dementia diagnosis |  |  |
| Median (range) | 72.4 (22.7-93.2) | 71.7 (43.8-79.6) |

As shown in **Figure 2**, hospitalisation for any infectious disease was associated with a 1·6- fold (95% CI 1·47–1·72) increased risk of subsequent dementia. The risk associated with any bacterial infection was of similar magnitude. When including only dementia cases occurring more than 10 years after the infection, these hazard ratios diminished and were modest, but remained statistically significant (1·26, 95% CI 1·14–1·40 for any infectious disease and 1·27, 95% CI 1·14–1·41 for any bacterial infection). The associations of bacterial infections did not differ by the invasiveness of the infection (invasive vs localised infections) or extracellular vs intracellular lifestyle of the causative microorganism. In contrast, Gram-negative bacterial infections were robustly associated with dementia even in the long-term, with a 1·6-fold increased risk of dementia in the full follow up and a 1·7- fold increased risk when dementia occurred at least 10 years after the infection. In addition, the excess risk associated with Gram-negative bacterial infections remained significant (HR 1·81, 95% CI 1·37–2·39) after excluding those with meningococcal, spirochaetal, and leptospiral infections, which are known for their ability to affect the brain.^18,19^

**Figure 2.**
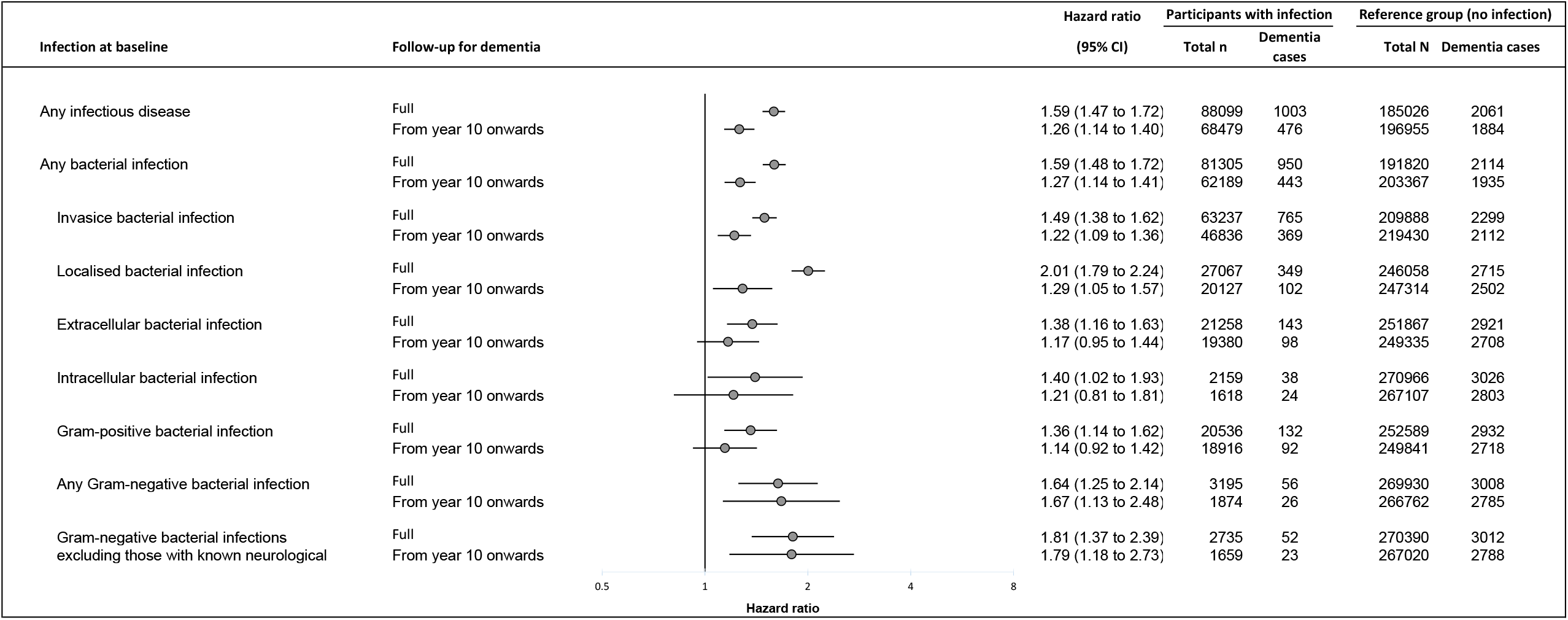
The risk of dementia associated with any hospital-treated infectious disease, any bacterial infection, and subgroups of bacterial infections in primary analysis. CI, confidence interval. Hazard ratios are adjusted for sex and socioeconomic status, and age is the time scale. *Infections caused by meningococci, spirochaetes, and leptospira excluded.

As shown in Figure 3, all viral infections combined were associated with a 1·6-fold (95% CI 1·32–2·00) increased risk of dementia, but the risk differed by subtype of viral infection. Acute viral infections were related to a 1·5-fold (95% CI 1·12–1·87) and herpesvirus infections and other potentially persistent viral infections (including hepatitis B and C, and HIV) to a 2-fold (95% CI 1·31–2·93 and 1·25–3·72, respectively) excess dementia risk. Only herpesvirus infections were robustly associated with dementia regardless of the follow-up duration (full or from year 10 onwards).

**Figure 3.**
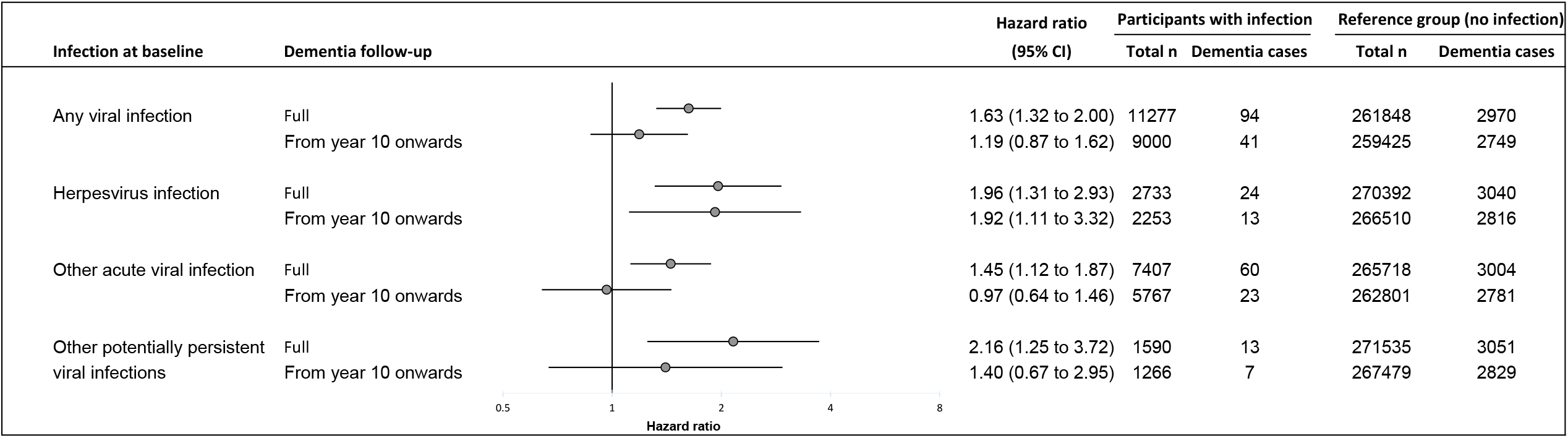
The risk of dementia associated with any hospital-treated viral infection and subgroups of viral infections in primary analysis. Legend: CI, confidence interval. Hazard ratios are adjusted for sex and socioeconomic status, and age is the time scale.

In sensitivity analyses (**Table 2**), the associations of gram-negative bacterial and herpesvirus infections with dementia were robust to exclusions of participants with hypertension, diabetes, ischaemic heart disease, cerebrovascular disease, and Parkinson’s disease and adjustment for lifestyle factors. In addition, these associations were replicable in the UK Biobank, in which 2375 dementia cases were recorded in 4 364 509 person-years at risk (median follow-up of 9·0 years). Gram-negative bacterial and herpesvirus infections were robustly related to all-cause dementia (both early- and late-onset) with stronger associations observed for vascular dementia than Alzheimer’s disease.

**Table 2.**
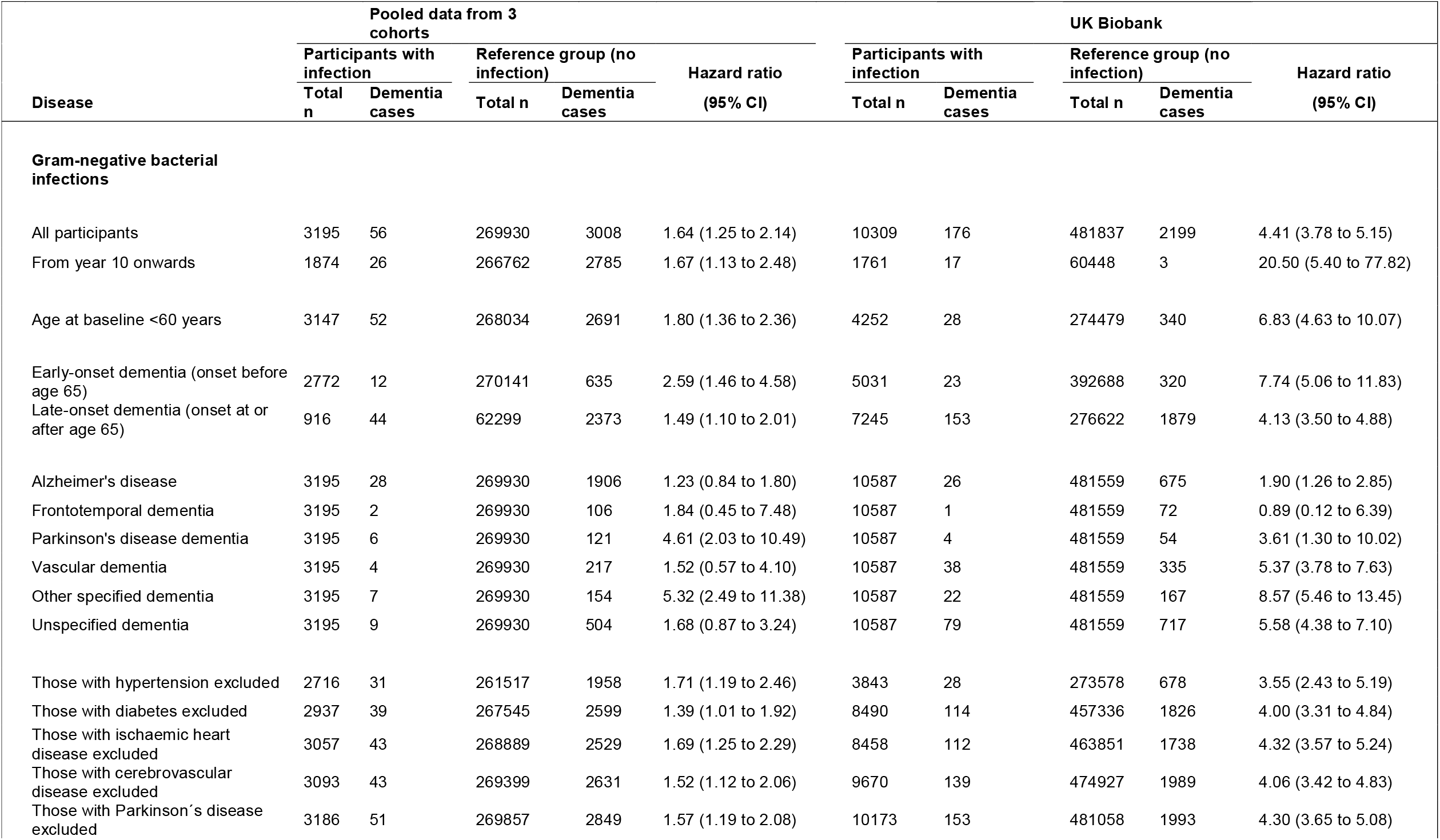
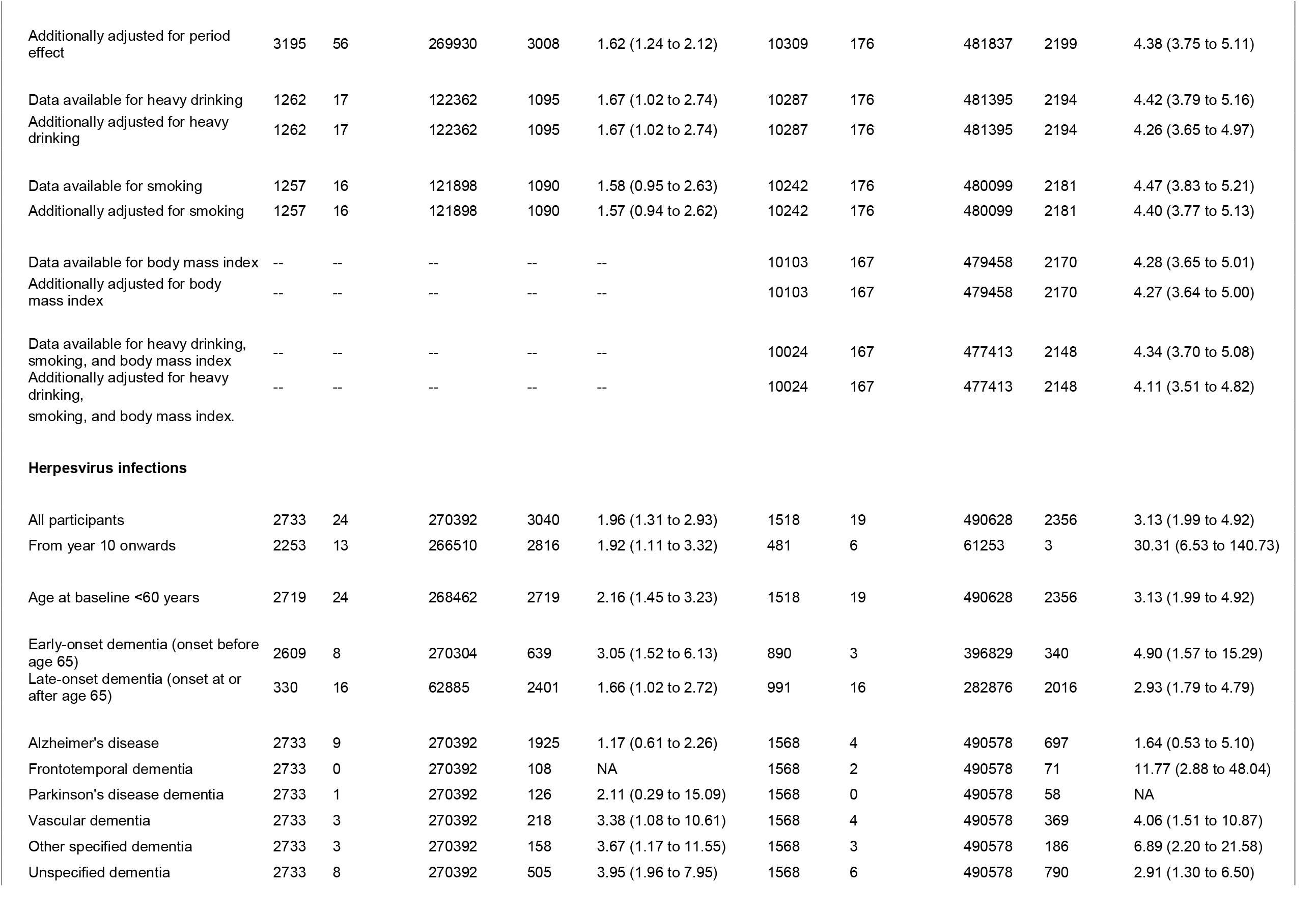
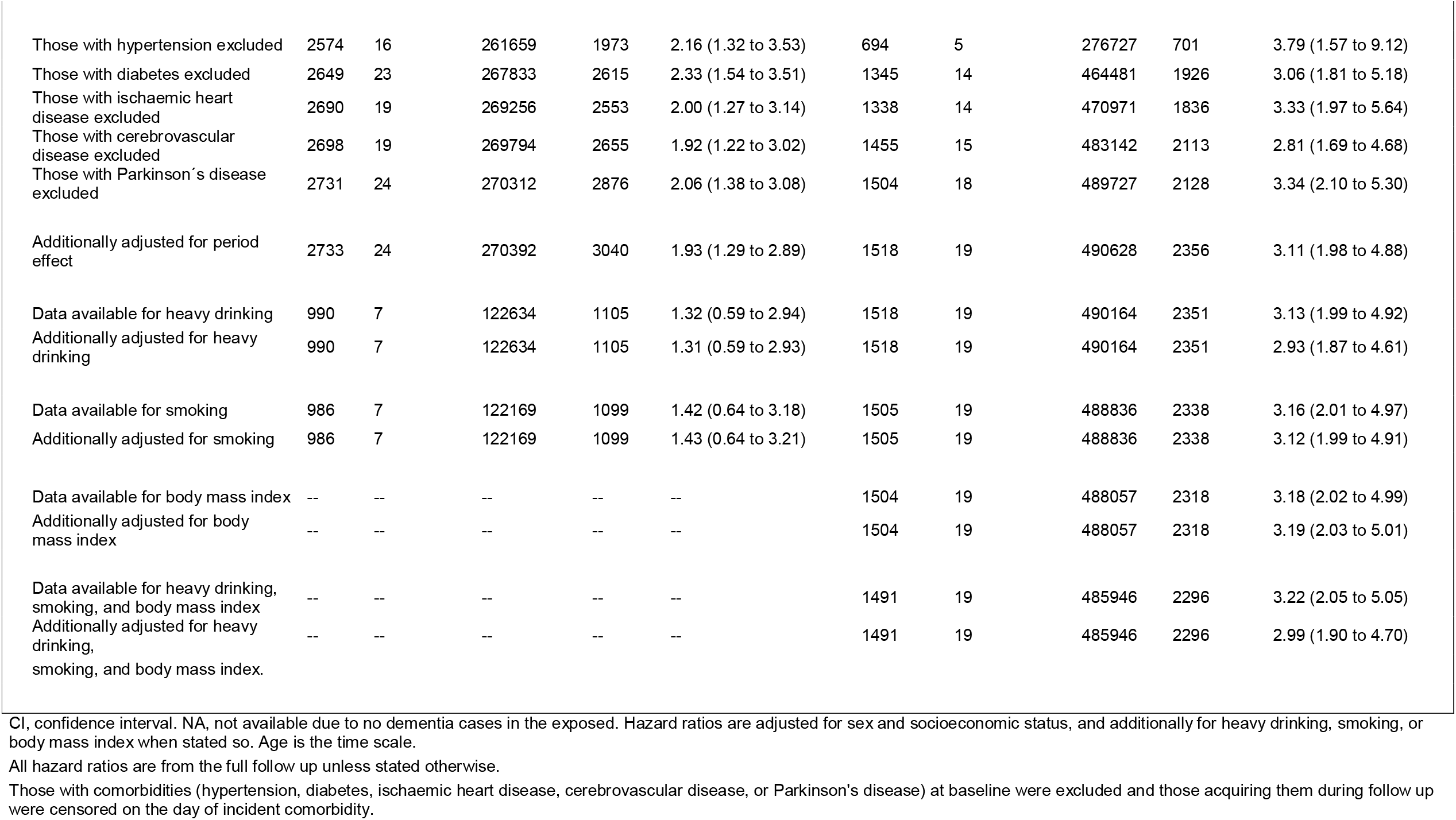
Associations of Gram-negative bacterial infections and herpesvirus infections with the risk of subsequent dementia in relation to follow-ups, types of dementia, adjustments and exclusions of participant groups in primary and replication cohorts.

| Disease | Pooled data from 3 cohorts |  |  |  |  | UK Biobank |  |  |  |  |
| --- | --- | --- | --- | --- | --- | --- | --- | --- | --- | --- |
|  | Participants with infection |  | Reference group (no infection) |  | Hazard ratio<br>(95% CI) | Participants with infection |  | Reference group (no infection) |  | Hazard ratio<br>(95% CI) |
|  | Total n | Dementia cases | Total n | Dementia cases |  | Total n | Dementia cases | Total n | Dementia cases |  |
| Gram-negative bacterial infections |  |  |  |  |  |  |  |  |  |  |
| All participants | 3195 | 56 | 269930 | 3008 | 1.64 (1.25 to 2.14) | 10309 | 176 | 481837 | 2199 | 4.41 (3.78 to 5.15) |
| From year 10 onwards | 1874 | 26 | 266762 | 2785 | 1.67 (1.13 to 2.48) | 1761 | 17 | 60448 | 3 | 20.50 (5.40 to 77.82) |
| Age at baseline <60 years | 3147 | 52 | 268034 | 2691 | 1.80 (1.36 to 2.36) | 4252 | 28 | 274479 | 340 | 6.83 (4.63 to 10.07) |
| Early-onset dementia (onset before age 65) | 2772 | 12 | 270141 | 635 | 2.59 (1.46 to 4.58) | 5031 | 23 | 392688 | 320 | 7.74 (5.06 to 11.83) |
| Late-onset dementia (onset at or after age 65) | 916 | 44 | 62299 | 2373 | 1.49 (1.10 to 2.01) | 7245 | 153 | 276622 | 1879 | 4.13 (3.50 to 4.88) |
| Alzheimer's disease | 3195 | 28 | 269930 | 1906 | 1.23 (0.84 to 1.80) | 10587 | 26 | 481559 | 675 | 1.90 (1.26 to 2.85) |
| Frontotemporal dementia | 3195 | 2 | 269930 | 106 | 1.84 (0.45 to 7.48) | 10587 | 1 | 481559 | 72 | 0.89 (0.12 to 6.39) |
| Parkinson's disease dementia | 3195 | 6 | 269930 | 121 | 4.61 (2.03 to 10.49) | 10587 | 4 | 481559 | 54 | 3.61 (1.30 to 10.02) |
| Vascular dementia | 3195 | 4 | 269930 | 217 | 1.52 (0.57 to 4.10) | 10587 | 38 | 481559 | 335 | 5.37 (3.78 to 7.63) |
| Other specified dementia | 3195 | 7 | 269930 | 154 | 5.32 (2.49 to 11.38) | 10587 | 22 | 481559 | 167 | 8.57 (5.46 to 13.45) |
| Unspecified dementia | 3195 | 9 | 269930 | 504 | 1.68 (0.87 to 3.24) | 10587 | 79 | 481559 | 717 | 5.58 (4.38 to 7.10) |
| Those with hypertension excluded | 2716 | 31 | 261517 | 1958 | 1.71 (1.19 to 2.46) | 3843 | 28 | 273578 | 678 | 3.55 (2.43 to 5.19) |
| Those with diabetes excluded | 2937 | 39 | 267545 | 2599 | 1.39 (1.01 to 1.92) | 8490 | 114 | 457336 | 1826 | 4.00 (3.31 to 4.84) |
| Those with ischaemic heart disease excluded | 3057 | 43 | 268889 | 2529 | 1.69 (1.25 to 2.29) | 8458 | 112 | 463851 | 1738 | 4.32 (3.57 to 5.24) |
| Those with cerebrovascular disease excluded | 3093 | 43 | 269399 | 2631 | 1.52 (1.12 to 2.06) | 9670 | 139 | 474927 | 1989 | 4.06 (3.42 to 4.83) |
| Those with Parkinson's disease excluded | 3186 | 51 | 269857 | 2849 | 1.57 (1.19 to 2.08) | 10173 | 153 | 481058 | 1993 | 4.30 (3.65 to 5.08) |
| Additionally adjusted for period effect | 3195 | 56 | 269930 | 3008 | 1.62 (1.24 to 2.12) | 10309 | 176 | 481837 | 2199 | 4.38 (3.75 to 5.11) |
| Data available for heavy drinking | 1262 | 17 | 122362 | 1095 | 1.67 (1.02 to 2.74) | 10287 | 176 | 481395 | 2194 | 4.42 (3.79 to 5.16) |
| Additionally adjusted for heavy drinking | 1262 | 17 | 122362 | 1095 | 1.67 (1.02 to 2.74) | 10287 | 176 | 481395 | 2194 | 4.26 (3.65 to 4.97) |
| Data available for smoking | 1257 | 16 | 121898 | 1090 | 1.58 (0.95 to 2.63) | 10242 | 176 | 480099 | 2181 | 4.47 (3.83 to 5.21) |
| Additionally adjusted for smoking | 1257 | 16 | 121898 | 1090 | 1.57 (0.94 to 2.62) | 10242 | 176 | 480099 | 2181 | 4.40 (3.77 to 5.13) |
| Data available for body mass index | -- | -- | -- | -- | -- | 10103 | 167 | 479458 | 2170 | 4.28 (3.65 to 5.01) |
| Additionally adjusted for body mass index | -- | -- | -- | -- | -- | 10103 | 167 | 479458 | 2170 | 4.27 (3.64 to 5.00) |
| Data available for heavy drinking, smoking, and body mass index | -- | -- | -- | -- | -- | 10024 | 167 | 477413 | 2148 | 4.34 (3.70 to 5.08) |
| Additionally adjusted for heavy drinking, smoking, and body mass index. | -- | -- | -- | -- | -- | 10024 | 167 | 477413 | 2148 | 4.11 (3.51 to 4.82) |
| **Herpesvirus infections** |  |  |  |  |  |  |  |  |  |  |
| All participants | 2733 | 24 | 270392 | 3040 | 1.96 (1.31 to 2.93) | 1518 | 19 | 490628 | 2356 | 3.13 (1.99 to 4.92) |
| From year 10 onwards | 2253 | 13 | 266510 | 2816 | 1.92 (1.11 to 3.32) | 481 | 6 | 61253 | 3 | 30.31 (6.53 to 140.73) |
| Age at baseline <60 years | 2719 | 24 | 268462 | 2719 | 2.16 (1.45 to 3.23) | 1518 | 19 | 490628 | 2356 | 3.13 (1.99 to 4.92) |
| Early-onset dementia (onset before age 65) | 2609 | 8 | 270304 | 639 | 3.05 (1.52 to 6.13) | 890 | 3 | 396829 | 340 | 4.90 (1.57 to 15.29) |
| Late-onset dementia (onset at or after age 65) | 330 | 16 | 62885 | 2401 | 1.66 (1.02 to 2.72) | 991 | 16 | 282876 | 2016 | 2.93 (1.79 to 4.79) |
| Alzheimer's disease | 2733 | 9 | 270392 | 1925 | 1.17 (0.61 to 2.26) | 1568 | 4 | 490578 | 697 | 1.64 (0.53 to 5.10) |
| Frontotemporal dementia | 2733 | 0 | 270392 | 108 | NA | 1568 | 2 | 490578 | 71 | 11.77 (2.88 to 48.04) |
| Parkinson's disease dementia | 2733 | 1 | 270392 | 126 | 2.11 (0.29 to 15.09) | 1568 | 0 | 490578 | 58 | NA |
| Vascular dementia | 2733 | 3 | 270392 | 218 | 3.38 (1.08 to 10.61) | 1568 | 4 | 490578 | 369 | 4.06 (1.51 to 10.87) |
| Other specified dementia | 2733 | 3 | 270392 | 158 | 3.67 (1.17 to 11.55) | 1568 | 3 | 490578 | 186 | 6.89 (2.20 to 21.58) |
| Unspecified dementia | 2733 | 8 | 270392 | 505 | 3.95 (1.96 to 7.95) | 1568 | 6 | 490578 | 790 | 2.91 (1.30 to 6.50) |
| Those with hypertension excluded | 2574 | 16 | 261659 | 1973 | 2.16 (1.32 to 3.53) | 694 | 5 | 276727 | 701 | 3.79 (1.57 to 9.12) |
| Those with diabetes excluded | 2649 | 23 | 267833 | 2615 | 2.33 (1.54 to 3.51) | 1345 | 14 | 464481 | 1926 | 3.06 (1.81 to 5.18) |
| Those with ischaemic heart disease excluded | 2690 | 19 | 269256 | 2553 | 2.00 (1.27 to 3.14) | 1338 | 14 | 470971 | 1836 | 3.33 (1.97 to 5.64) |
| Those with cerebrovascular disease excluded | 2698 | 19 | 269794 | 2655 | 1.92 (1.22 to 3.02) | 1455 | 15 | 483142 | 2113 | 2.81 (1.69 to 4.68) |
| Those with Parkinson's disease excluded | 2731 | 24 | 270312 | 2876 | 2.06 (1.38 to 3.08) | 1504 | 18 | 489727 | 2128 | 3.34 (2.10 to 5.30) |
| Additionally adjusted for period effect | 2733 | 24 | 270392 | 3040 | 1.93 (1.29 to 2.89) | 1518 | 19 | 490628 | 2356 | 3.11 (1.98 to 4.88) |
| Data available for heavy drinking | 990 | 7 | 122634 | 1105 | 1.32 (0.59 to 2.94) | 1518 | 19 | 490164 | 2351 | 3.13 (1.99 to 4.92) |
| Additionally adjusted for heavy drinking | 990 | 7 | 122634 | 1105 | 1.31 (0.59 to 2.93) | 1518 | 19 | 490164 | 2351 | 2.93 (1.87 to 4.61) |
| Data available for smoking | 986 | 7 | 122169 | 1099 | 1.42 (0.64 to 3.18) | 1505 | 19 | 488836 | 2338 | 3.16 (2.01 to 4.97) |
| Additionally adjusted for smoking | 986 | 7 | 122169 | 1099 | 1.43 (0.64 to 3.21) | 1505 | 19 | 488836 | 2338 | 3.12 (1.99 to 4.91) |
| Data available for body mass index | -- | -- | -- | -- | -- | 1504 | 19 | 488057 | 2318 | 3.18 (2.02 to 4.99) |
| Additionally adjusted for body mass index | -- | -- | -- | -- | -- | 1504 | 19 | 488057 | 2318 | 3.19 (2.03 to 5.01) |
| Data available for heavy drinking, smoking, and body mass index | -- | -- | -- | -- | -- | 1491 | 19 | 485946 | 2296 | 3.22 (2.05 to 5.05) |
| Additionally adjusted for heavy drinking, smoking, and body mass index. | -- | -- | -- | -- | -- | 1491 | 19 | 485946 | 2296 | 2.99 (1.90 to 4.70) |

CI, confidence interval. NA, not available due to no dementia cases in the exposed. Hazard ratios are adjusted for sex and socioeconomic status, and additionally for heavy drinking, smoking, or body mass index when stated so. Age is the time scale.
All hazard ratios are from the full follow up unless stated otherwise.
Those with comorbidities (hypertension, diabetes, ischaemic heart disease, cerebrovascular disease, or Parkinson's disease) at baseline were excluded and those acquiring them during follow up were censored on the day of incident comorbidity.

In post-hoc analyses, we compared the distribution of different Gram-negative bacterial infections between incident cases and non-cases of dementia (appendix, pp. 78). Sepsis was more common in those who developed dementia (*P* = 0·02), but sepsis due to Gram-negative bacteria and other Gram-negative infections had identical associations with incident dementia (both hazard ratios 1·63).

## Discussion

We assessed dementia risk associated with 528 hospital-treated bacterial and viral infections in a pooled analysis of over 270 000 adults followed up for a median of 19 years. Gram-negative bacterial infections were related to a 1.6-fold and herpesvirus infections to a 2-fold increased risk of subsequent dementia both in the full follow up and when reverse causation was minimised by restricting analyses to infections that occurred more than 10 years before dementia onset. These associations were robust to adjustments for cardiovascular and other comorbidities and lifestyle-related factors. These associations were also replicated in an independent cohort, the UK Biobank, which includes 492 146 adults. In contrast, any associations of hospital-treated bacterial and viral infections in general or acute viral infections specifically were either modest or attenuated in analyses controlling for reverse causation and ascertainment biases.

To our knowledge, the robust long-term association between Gram-negative bacterial infections and dementia is a novel finding, although it is not implausible considering the extant literature. Some Gram-negative bacteria are known for their ability to affect the brain. For example, *Neisseria meningitidis* (meningococcus) is an important cause of bacterial meningitis^18^ and the spirochaete *Treponema pallidum* can cause neurosyphilis that may lead to dementia.^19^ The association between Gram-negative bacteria and dementia could not be explained by the specific contribution of meningococcal or spirochaetal infections. Rather, Gram-negative bacterial infections in general contributed to increased dementia risk, a finding consistent with earlier evidence for the role of inflammation in dementia.^1,8,9^

Recent animal models support specific mechanisms linking Gram-negative bacterial infections to dementia.^2^ Gram-negative bacteria produce lipopolysaccharides that are potent stimulators of the immune system.^25^ Lipopolysaccharides can prime microglia to a proinflammatory state that may persist long after the initial insult,^2^ potentially increasing the deposition of amyloid plaques, which characterise Alzheimer’s disease.^2^ We found that the relationship of Gram-negative bacterial infections was stronger to vascular dementia than Alzheimer’s disease suggesting that mechanisms beyond amyloid deposition are also likely to contribute to dementia risk.

Herpesvirus infections were another group of infectious diseases with a robust long-term association with dementia. Our findings are consistent with a recent systematic review that found a suggestive association between a recent herpesvirus infection or reactivation and dementia.^16^ No association was observed for herpesvirus seropositivity, an inclusive exposure definition that also captures cases with asymptomatic infection. It is possible that excess dementia risk is attributable to severe symptomatic primary infection or viral reactivation, indicated by hospitalisations, rather than to mild herpesvirus infections. Supporting the herpesvirus-dementia association, other studies have found increased herpesvirus activity in the brains of people with Alzheimer’s disease,^3^ that anti-herpetic medications might reduce the risk of dementia in those with herpesvirus infections,^26^ and that protection against infectious agents, such as herpesviruses, may even be one of the biological functions of β-amyloid.^4^ However, similarly with Gram-negative bacterial infections, herpesvirus infections were more strongly associated with vascular dementia than Alzheimer’s disease. Further mechanistic research is needed to test the hypothesis that these infections may affect in particular the risk of developing vascular dementia, as the ascertainment of dementia subtypes from health records in our study is uncertain and clinical dementia is a syndrome that often involves multiple underlying neuropathologies.^27^

The associations of Gram-positive and other bacterial infections and non-herpetic viral infections with long-term dementia risk were, if anything, modest. This suggests that the observed short-term associations between these infections and dementia may be inflated by confounding or reverse causation due to factors such as declining host resistance during preclinical dementia.^11^ In two cohort studies not accounting for these biases, any hospital-treated infection was associated with a 2-fold increased risk of subsequent dementia.^11,13^ A third cohort study that excluded the first 2 years of follow-up to reduce bias^12^ revealed a weak association with a modest hazard ratio of 1·2, which is similar to the 1·2- to 1·3-fold increased risk of dementia in our bias-corrected analyses. We found no robust association between acute viral infections and dementia, consistent with an earlier study that reported no association between influenza and dementia.^28^

Hospital-treated invasive bacterial infections were not associated with a higher risk of dementia than localised bacterial infections and at least two earlier studies have reported similar findings.^11,12^ We assessed hospital-treated infections, which are likely to be relatively severe irrespective of whether they are considered invasive or localised. Thus, if there is a biological pathway through which infectious diseases contribute to the development of dementia (possibly acting through innate immune memory or some other inflammatory mechanism),^2^ most hospital-treated infections may be sufficiently potent to activate that pathway at least to some extent.

This study has some important strengths. We systematically investigated hospitalisations for different types of infectious diseases and were able to replicate the main findings in a large independent cohort. We used detailed discharge records to identify infections at the level of causative microorganism. In contrast to earlier analyses, we examined long-term associations that were less likely to be attributable to reverse causation or ascertainment bias. As our disease ascertainment was based on nationwide register data rather than participation in a clinical examination, our follow-up was not dependent on a person’s active participation in study examinations during follow up and thus was virtually complete. The main limitations of this study include the lack of a uniform investigator-led examination of all participants to identify cases of milder infections; residual confounding by frailty, undiagnosed comorbidities, medications (such as antibiotics),^30^ and ascertainment of dementia based on linkage to electronic health records, which is likely to miss milder cases of dementia^30^ and lacks detailed diagnosis of underlying neuropathology.

In conclusion, hospital-treated Gram-negative bacterial infections and herpesvirus infections were robustly associated with a moderately increased risk of all-cause dementia and early and late-onset dementia. These associations persisted in the long-term and were not attributable to co-morbid cardiometabolic conditions, Parkinson’s disease, lifestyle factors, or obesity. Further mechanistic and molecular research is warranted to clarify the potential role of Gram-negative bacterial infections and herpesviruses in neurodegeneration and the aetiology of dementia.

## Contributors

PNS and MK developed the study idea. PNS, NH, and MK designed the study. PNS and CH analysed the data. PNS conducted the literature review and wrote the first draft of the manuscript. MK obtained funding for the study. JV, SS, AV, and MK contributed to data acquisition. PNS, NH, JVL, CH, JV, ME, SS, AK, STN, JP, TES, and MK contributed to the study conception or design or analysis or interpretation of the data. All authors contributed to critical revision of the manuscript for important intellectual content and approved the submitted manuscript. PNS, JP, and MK had full access to all primary data and CH to all replication data in the study and take responsibility for the integrity of the data and the accuracy of the data analysis. PNS is the guarantor.

## Data Availability

The data of the primary cohorts are not publicly available due to privacy restrictions. The data of the replication cohort (UK Biobank) are open to researchers: https://www.ukbiobank.ac.uk/researchers/.

## Acknowledgements

The study was supported by grants from NordForsk, the Academy of Finland (311492, 310591), the Helsinki Institute of Life Science, the US National Institute on Aging (R01AG056477), and the UK Medical Research Council (MRC S011676/1, R024227/1). This research has been conducted using the UK Biobank Resource under Application Number 14801.

